# Beyond single-slope Mendelian randomization: structural representation of genetic heterogeneity in joint effect space

**DOI:** 10.64898/2026.03.12.26348288

**Authors:** Hongdong Hao, Dian Chen, Cheng Qian, Xuanyi Zhou, Hao Huang, Jiayu Zuo, Guanlin Wang, Xi Peng, Hai-Xin Liu

## Abstract

Causal effects in complex traits are typically represented by a single linear slope. While conventional Mendelian randomization (MR) provides efficient scalar estimates, projection-based summaries do not explicitly capture structural organisation in joint effect space under genetic heterogeneity. We introduce MR-UBRA (Mendelian randomization–Unified Bayesian Risk Architecture), a probabilistic framework that decomposes instrumental variants into genetic risk fragments (GRFs) and quantifies extreme deviations using tail-risk metrics defined on the standardised residual magnitude |e|. MR-UBRA preserves the classical MR estimand while offering a structurally resolved representation of genetic heterogeneity.

Across stroke subtypes, AF→CES, smoking→lung cancer, and BMI→T2D, effect-space distributions exhibit reproducible asymmetry, amplitude stratification, and multi-modal structure. MR-UBRA resolves component-level organisation, separating tail-dominant contributions from the causal core while maintaining consistency with the classical MR estimand.

Simulations and boundary regimes demonstrate adaptive model complexity: MR-UBRA selects K>1 when multi-component structure is present and collapses to K=1 under homogeneous conditions, avoiding spurious stratification.

These results support viewing causal effects in complex traits as structured distributions in joint effect space, enhancing causal representation without altering the MR estimand.

**Graphical Abstract:** Mendelian randomization (MR) typically represents causal effects with a single linear slope. Under genetic heterogeneity, instrumental effects in joint (βX, βY) space may exhibit multi-component structure and amplitude stratification that cannot be captured by a scalar summary. MR-UBRA fits a standard error–weighted mixture model to decompose instruments into genetic risk fragments (GRFs), estimates GRF-specific effects using posterior-weighted soft-IVW, and quantifies extreme deviations through tail-risk metrics (VaR/CVaR). Across empirical analyses and boundary regimes, MR-UBRA adapts model complexity (K) to structural signal, collapsing to K=1 under homogeneous conditions.

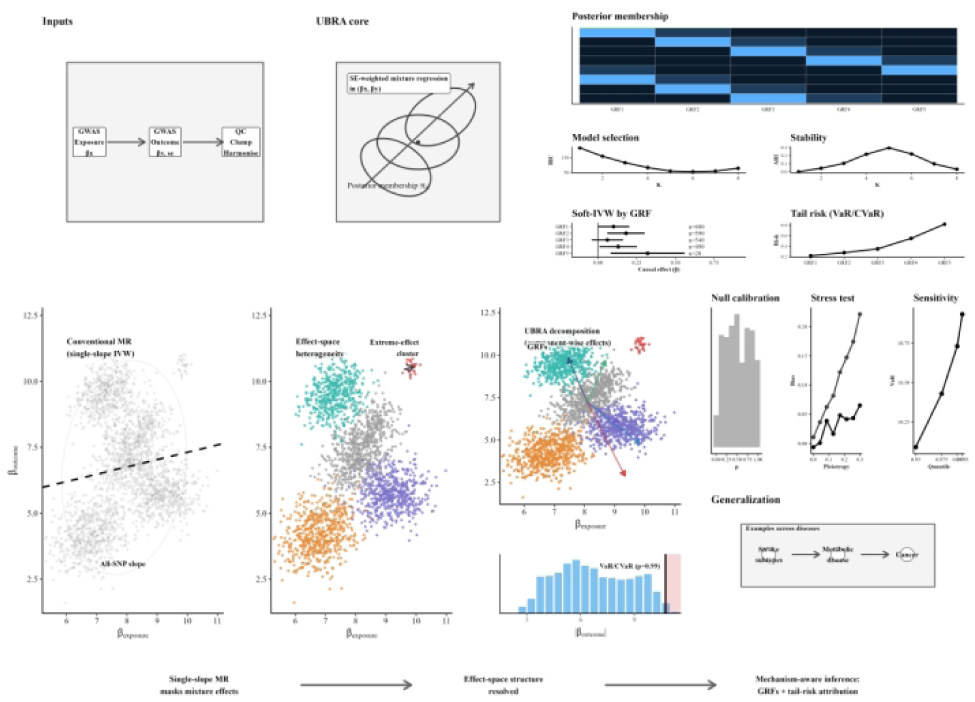

## 1 Introduction

### 1.1 Statistical Background of Single-Slope Causal Representation in Complex Traits

Single-slope Mendelian randomization (MR) is a projection-based representation of causal effects in joint effect space[1, 2]. When effect structures are homogeneous and symmetric, this approach efficiently captures exposure–outcome relationships[3, 4]. However, as genetic architectures in complex traits exhibit increasing heterogeneity[5, 6], instrumental effects in joint effect space may exhibit multi-component structure and amplitude stratification, which are not captured by a single slope. A single-slope representation retains directional inference while obscuring underlying structural organisation[7].

### 1.2 A Structural Representational Gap in Current Mendelian Randomization Frameworks

Current MR frameworks remain projection-centric, even with heterogeneity-aware estimators such as MR-Egger, weighted median, and MR-Clust[8, 9]. While these methods improve bias control and detect opposing directional effects, they summarise causal effects with a single slope, offering limited representation of structure within joint effect space[7, 10]. Heterogeneity is treated as residual variation under projection-based summaries[11, 12]. This gap motivates an explicit characterisation of genetic effects in joint effect space[13].

### 1.3 Empirical Evidence of Tail-Dominant Structure in Joint Effect Space

Genetic effects in joint (βX, βY) effect space frequently exhibit distributional asymmetry, heavy-tailed deviation, and amplitude stratification[14, 15]. Effect distributions show multi-modal structure, with a small subset of high-magnitude variants contributing disproportionately to overall statistics[16, 17]. Observed heterogeneity in effect space reflects structured organisation rather than random variation[18, 19].

### 1.4 An Effect-Space Perspective Inspired by Tail-Risk Metrics

We use tail-risk metrics as distributional descriptors of extreme deviation concentration[20, 21]. Tail-risk metrics, defined on standardised residual magnitude |e|, quantify tail concentration and deviation localisation[21, 22]. These metrics provide a structured, statistical representation of genetic heterogeneity in complex traits[23, 24].

### 1.5 Study Contributions and Scope

We define causal effects in joint effect space as a structured distribution composed of multiple genetic risk factors (GRFs)[23, 25]. We do not redefine the classical MR estimand, but provide a higher-resolution statistical representation of genetic heterogeneity[2]. MR-UBRA models the overall effect as a multi-component distribution, preserving directional inference while decomposing contributions across amplitude strata[24, 26]. This framework is applied to diverse complex trait relationships, offering a structured representation of genetic effects across heterogeneous disease contexts.

## 2 Methods

### 2.1 MR-UBRA (UBRA-2D) Algorithmic Framework

#### 2.1.1 Model Overview

MR-UBRA conceptualizes causal effects as a structured distribution embedded in a two-dimensional joint effect space[27]. For each genetic variant, we represent its associations with both exposure and outcome as a vector z_j_ = (β^_Xj_, β^_Yj_), thereby preserving the geometric relationship between these estimates rather than collapsing them into a univariate summary[28].

The framework decomposes the genetic architecture into two fundamental components: a causal backbone that captures the primary directional signal, and structured deviations that reveal reproducible heterogeneity. The backbone is formally defined as the zero-intercept weighted projection slope, representing the consensus causal effect after accounting for sampling variability. Structured deviations—particularly those exhibiting large magnitudes (|e_j_|)—are explicitly modelled to identify distinct genetic regulatory foci (GRFs) that may reflect horizontal pleiotropy, population stratification, or biologically meaningful pathway-specific effects.

Rather than reducing the data to a single global estimate, this formulation preserves the full geometric structure of the effect-size distribution. Deviation patterns are decomposed using a probabilistic mixture model with soft allocation, enabling variants to contribute fractionally to multiple GRFs based on their posterior membership probabilities. The framework generates a comprehensive suite of outputs: global backbone estimates, GRF-specific causal effects, tail-risk statistics quantifying extreme deviations, leave-one-GRF-out influence diagnostics, allocation uncertainty metrics, structural stability measures, and empirically calibrated significance assessments. All results are systematically exported as standardized tables, publication-ready figures (PNG/PDF formats), and complete runtime metadata to ensure full reproducibility.

#### 2.1.2 Effect-Space Representation and Weighting

For each SNP j, let β^_Xj_ and β^_Yj_ denote the estimated exposure and outcome effects, respectively. These quantities define the joint effect vector

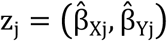

which serves as the fundamental unit of analysis.

To appropriately account for differential estimation precision, we assign a weight w_j_ to each SNP. When outcome standard errors are available, precision-based weighting is employed using the inverse variance:

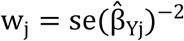

In cases where standard errors are unavailable, uniform weighting (w_j_ = 1) is applied as a conservative alternative.

Quality control proceeds through sequential filtering: SNPs must have finite values for both effect estimates and positive weights to be retained. An optional lower bound on |β^_Xj_| may be imposed to enhance numerical stability in the presence of very weak instruments[29]. If fewer than 50 SNPs survive these filters, the structural decomposition is aborted and only the global backbone estimate is reported—a contingency clearly documented as “SKIPPED” in the runtime metadata to maintain analytical transparency.

#### 2.1.3 Probabilistic Mixture Decomposition and Soft Allocation

To systematically characterize heterogeneity in the joint effect space, we model the empirical distribution of SNP effect vectors using a K -component two-dimensional Gaussian mixture model[30]:

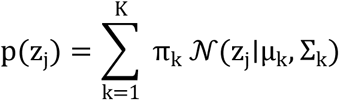

Here, π_k_ represents the mixing proportion for component k, while μ_k_ and Σ_k_denote its mean vector and covariance matrix, respectively. During model fitting, effects are centered and standardized to ensure numerical stability, with all final estimates transformed back to the original scale for interpretability.

A key feature of this approach is its probabilistic treatment of cluster membership. Rather than imposing hard assignments, we compute posterior membership probabilities:

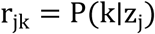

which quantify the degree of belongingness of each SNP to each GRF. This soft allocation framework naturally accommodates variants that may exhibit intermediate characteristics or shared influences across multiple biological pathways.

The uncertainty inherent in these assignments is characterized through multiple complementary metrics: the maximum posterior probability maxr_jk_ indicates assignment confidence; the posterior entropy – Σ_k_ r_jk_ logr_jk_ captures the overall dispersion of membership probabilities; and the gap between the two largest posterior probabilities provides insight into the distinctiveness of the primary assignment. Together, these measures offer a nuanced assessment of allocation certainty at the individual variant level.

#### 2.1.4 Model Complexity and Structural Stability

Selecting the appropriate number of mixture components K represents a critical model choice that balances descriptive adequacy against parsimony. We conduct a systematic search over a predefined range (K ∈ {1, …, K_max_}), evaluating each candidate through the dual lenses of goodness-of-fit and structural stability.

Goodness-of-fit is assessed using the Bayesian Information Criterion (BIC), which penalizes model complexity while rewarding improvements in likelihood[30, 31]. However, recognizing that statistical fit alone may select overly complex models with poor replicability, we additionally evaluate structural stability through a robust subsampling procedure[32]. For each candidate K, we repeatedly refit the model on random subsamples of the data and compute the adjusted Rand index (ARI) between the resulting cluster assignments[33]. High ARI values indicate that the discovered structure is reproducible rather than driven by idiosyncratic features of the full sample.

A model with larger K is accepted only when it satisfies three criteria simultaneously: (1) meaningful improvement in BIC (or optimal ranking among candidates), (2) stable cluster assignments under subsampling as evidenced by high ARI, and (3) absence of degenerate components characterized by near-zero mixture weights or singular covariance matrices. To ensure complete transparency and reproducibility, we record all candidate K trajectories, subsampling iterations, random seeds, and final selection criteria in the runtime metadata.

#### 2.1.5 Causal Backbone Estimation and Standardised Residuals

The global causal backbone—representing the consensus effect after accounting for heterogeneity—is estimated as a precision-weighted projection slope:

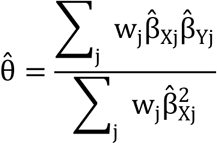

This estimator corresponds to the inverse-variance weighted (IVW) slope under the assumption of no pleiotropy, and serves as a natural benchmark against which individual variant deviations can be assessed.

To quantify departures from this backbone, we define standardized residuals:

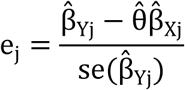

where the denominator defaults to unity when standard errors are unavailable. These residuals capture the extent to which each SNP’s outcome association deviates from the prediction implied by its exposure association and the global backbone. The absolute residual |e_j_| provides a direct measure of deviation magnitude, with large values indicating variants whose effects are poorly explained by the consensus causal model—potentially reflecting horizontal pleiotropy, measurement error, or genuine biological heterogeneity[1].

#### 2.1.6 Component Effects, Tail Risk, and Influence

Beyond global inference, we characterize effect heterogeneity at the GRF level. Component-specific causal effects are estimated by weighting SNP contributions according to their posterior membership probabilities:

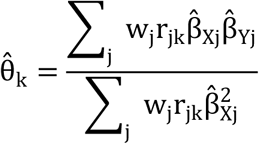

These GRF-level estimates reveal how the exposure-outcome relationship may differ across distinct genetic regulatory contexts.

To systematically characterize extreme deviations, we examine the upper tail of the absolute residual distribution. Value at Risk (VaR_ρ_) and Conditional Value at Risk (CVaR_ρ_) are computed empirically, with ρ = 0.99 as the default threshold unless otherwise specified[34]. These metrics quantify, respectively, the magnitude above which the largest 1% of absolute deviations fall, and the expected value of deviations exceeding that threshold.

For each GRF, we introduce the concept of exceedance energy—a measure of the component’s contribution to tail risk:

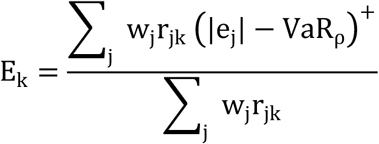

where (x)^+^ = max(x, 0). This quantity captures the average excess deviation among variants assigned to component k, weighted by their membership probabilities, providing insight into which GRFs are primarily responsible for extreme outliers.

The influence of each GRF on the global backbone is assessed through a leave-one-component-out analysis:

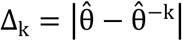

where θ^^-k^ is recomputed after setting r_jk_ = 0 for all SNPs in component k and re-estimating the backbone using the remaining weighted contributions. Large Δ_k_ values indicate GRFs that exert substantial leverage on the global estimate, warranting careful scrutiny.

#### 2.1.7 Empirical Calibration

To establish statistical significance for our heterogeneity metrics, we construct an empirical null distribution through residual permutation. This procedure preserves the marginal scale and weight structure of the original data while breaking any systematic relationship between residuals and component assignments. Under each permutation, we recompute VaR_ρ_, CVaR_ρ_, E_k_, and Δ_k_, thereby generating null distributions that reflect what would be expected under exchangeability.

Empirical p-values are calculated as the proportion of null statistics greater than or equal to the observed value. To account for multiple testing across GRFs and metrics, we apply the Benjamini–Hochberg procedure to control the false discovery rate, ensuring that reported findings are robust to multiplicity concerns[35].

#### 2.1.8 Diagnostics and Sensitivity

Comprehensive diagnostic assessment accompanies all primary analyses. Structural stability is evaluated through subsampling-based adjusted Rand indices and sensitivity to hyperparameter perturbations. Statistical significance of both stability metrics and component influence measures is assessed using the empirical calibration framework described above.

Within each identified GRF, we further characterize heterogeneity and potential pleiotropy using conventional meta-analytic tools: Cochran’s Q statistic quantifies residual heterogeneity beyond that explained by the component-level effect; I^2^ expresses this heterogeneity as a proportion of total variability; and the MR-Egger intercept tests for directional pleiotropy concentrated within the GRF[36, 37]. All sensitivity analyses are conducted under identical parameter locking and random seed specifications as the primary analysis, ensuring that comparisons are not confounded by stochastic variation.

### 2.2 Applied Analysis Workflow

#### 2.2.1 Data Sources

Ischemic stroke outcomes were derived from three complementary datasets within the MEGASTROKE consortium: MEGASTROKE.2.AIS.TRANS (all ischemic stroke), MEGASTROKE.3.LAS.TRANS (large artery stroke), and MEGASTROKE.4.CES.TRANS (cardioembolic stroke)[38]. Exposure and control trait genome-wide association study (GWAS) summary statistics were obtained from publicly available European and East Asian ancestry datasets.

#### 2.2.2 Instrument Selection, LD Control, and Harmonisation

Genetic instruments were selected using conventional genome-wide significance thresholds (P < 5 × 10^−8^)[39]. In sensitivity analyses where instrument counts were insufficient, pre-specified lower thresholds were applied and clearly documented. Linkage disequilibrium (LD) was addressed through approximate clumping within fixed physical windows, retaining the most significant SNP from each LD block[40].

Allele harmonization was performed systematically to ensure consistent effect orientation across datasets: strand flips were applied where required, palindromic SNPs with ambiguous allele frequencies were carefully evaluated, and variants with mismatched alleles or unresolvable ambiguities were excluded. The final harmonized dataset thus represents a conservative set of reliably aligned instruments[41].

#### 2.2.3 Quality Control and Low-SNP Contingency

Stringent quality control preceded all analyses: SNPs with non-finite effect estimates, implausibly extreme weights, or evidence of data corruption were removed. A dispersion-based filter on β^_X_ was optionally applied to exclude variants with unduly influential exposure effects. Crucially, analyses with fewer than 50 retained SNPs following QC were flagged and processed in backbone-only mode, with the full UBRA-2D decomposition automatically skipped and the contingency recorded as “SKIPPED” in runtime metadata. This threshold ensures that mixture modeling is attempted only when sufficient data exist to support reliable structural inference.

#### 2.2.4 Baseline Methods and Comparative Analyses

To contextualize UBRA-2D results within the existing MR literature, we computed conventional estimates using the same harmonized datasets: inverse-variance weighting (IVW), weighted median, and MR-Egger[8, 10]. Heterogeneity (Cochran’s Q) and pleiotropy (MR-Egger intercept) statistics were reported for each method. Additionally, we applied MR-Clust—a directional clustering approach—as a comparator for identifying distinct effect subgroups, using identical SNP inputs to ensure fair comparison.

#### 2.2.5 UBRA Decomposition and Cross-Task Synthesis

UBRA-2D was executed under unified parameter locking, with all specifications held constant across analyses to ensure comparability. The full pipeline encompassed K -selection, soft allocation, backbone estimation, GRF-level effect estimation, tail-risk computation, and leave-one-GRF-out influence analysis. For multi-task investigations (e.g., comparing atrial fibrillation to cardioembolic stroke), all analyses were processed within an integrated pipeline.

Standardized multi-panel visualizations were generated to facilitate cross-task synthesis: backbone comparison plots display GRF-level effects across phenotypes; tail-risk versus effect drift scatterplots reveal relationships between deviation magnitude and causal estimates; and geometric consistency maps illustrate the reproducibility of GRF structure across related outcomes.

### 2.3 Simulation and Boundary-Regime Validation

#### 2.3.1 Simulation Design

We conducted extensive simulations to evaluate UBRA-2D performance under controlled conditions. Data were generated under four scenarios: (1) Null (no causal effect, θ = 0); (2) Clean (homogeneous causal effect, single GRF); (3) Balanced (multiple GRFs with opposing directions); and (4) Multi-component (complex architectures with 3-5 distinct GRFs). For each scenario, independent datasets were repeatedly generated with fixed true θ and analyzed using both UBRA-2D and conventional IVW. Performance was assessed through bias, root mean square error (RMSE), and stability of K-selection across replications.

#### 2.3.2 Boundary-Regime Validation

To verify that UBRA-2D does not overfit when the true structure is simple, we specifically evaluated performance under single-component architectures. Under these conditions, we confirmed that the model consistently selects K = 1, that effect-space geometry reflects the expected homogeneity, that tail-risk metrics remain within null-expected ranges, and that influence statistics show no evidence of spurious influential GRFs. This validation ensures that the method defaults appropriately to simpler representations when the data do not support more complex structure.

### 2.4 Data and Code Availability

All analyses were conducted using publicly available GWAS summary statistics and MEGASTROKE consortium data, accessed under appropriate data use agreements. Complete analysis code is provided as executable scripts, including a unified runner, structured output generators, and a comprehensive reproducibility checklist. Runtime metadata systematically records all parameters, software versions, random seeds, and K-selection trajectories to facilitate independent verification[42].

The MR-UBRA implementation and all replication scripts for the atrial fibrillation to cardioembolic stroke analysis are openly available at: https://github.com/0808hao/MR-UBRA

No proprietary or custom hardware was required; all analyses were performed on standard computing infrastructure using open-source software environments.

## 3 Results

### 3.1 Systematic heterogeneity in genetic effect distributions and tail-risk structure across stroke subtypes

At the genome-wide significance threshold (P < 5 × 10⁻⁸), we compared genetic effect distributions for AIS, LAS, and CES within a unified joint effect-space framework. Clear geometric differences were observed across subtypes (Fig. 1A). AIS and LAS formed relatively compact and approximately symmetric point clouds, whereas CES displayed pronounced asymmetry with multi-modal structure, indicating organisation beyond a single central trend.

**Fig. 1.**
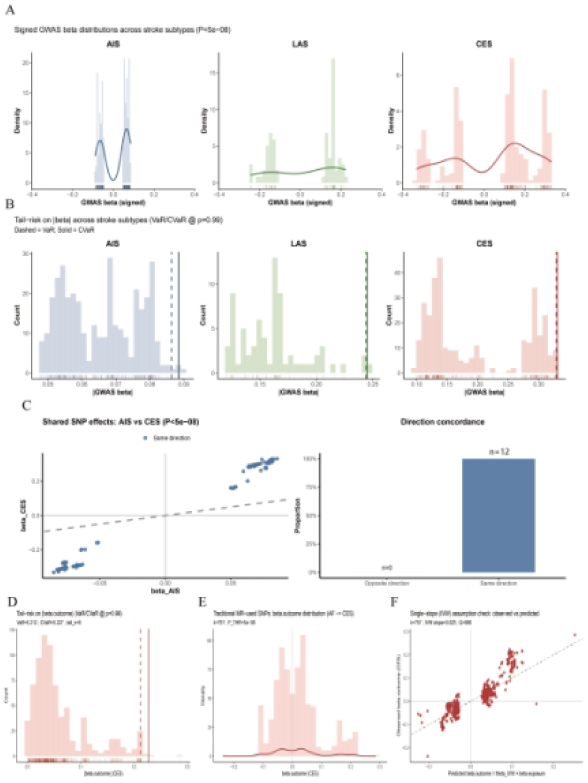
Effect-space geometry and tail-risk structure across stroke subtypes. A, Joint effect-space scatter of genome-wide significant variants (P < 5 × 10⁻⁸) for AIS, LAS, and CES. B, Tail-risk profiles computed on the standardised residual magnitude |e| (VaR/CVaR at ρ = 0.99). C, Effects of shared genome-wide significant variants between AIS and CES, showing direction concordance (n = 12). D, CES outcome-effect distribution highlighting a dense low-effect core and the VaR/CVaR-defined extreme region. E, Outcome-effect distribution among MR-selected variants in the AF → CES analysis. F, Single-slope (IVW) diagnostic: observed versus IVW-predicted outcome effects.

To quantify extreme deviation concentration, we characterised tail behaviour using tail-risk statistics computed on the standardised residual magnitude |e| (Fig. 1B). At ρ = 0.99, both VaR and CVaR were substantially higher for CES than for AIS and LAS, indicating stronger concentration of extreme deviations. In contrast, AIS and LAS exhibited smaller tail magnitudes, consistent with lighter or near-symmetric deviation distributions.

Shared genome-wide significant variants between AIS and CES showed largely concordant directions (Fig. 1C); however, directional agreement did not imply structural homogeneity. In CES, shared variants exhibited marked amplitude expansion, indicating that directionally aligned signals can occupy distinct intensity layers across subtypes. Consistently, the distribution of outcome effects in CES showed a dense low-effect core together with a limited set of variants residing in the VaR/CVaR-defined extreme region (Fig. 1D). Although few in number, these variants disproportionately shaped tail-risk statistics and overall distributional geometry.

Within instrument sets used for MR analysis, even after stringent significance filtering, IVW-included variants spanned a wide range of outcome effects (Fig. 1E). Comparing IVW-predicted versus observed outcome effects revealed systematic deviation, most evident in high-amplitude regions (Fig. 1F), indicating that a single-slope summary cannot simultaneously capture the low-effect bulk and the extreme tail structure.

Together, these results demonstrate stable and reproducible structural differences across stroke subtypes, with CES exhibiting the strongest asymmetry and tail extension. Under such multi-component organisation, the association is not fully captured by a single average slope.

### 3.2 Effect-space decomposition reveals tail-dominant structure in the AF→CES association

In the AF→CES analysis, 89 instrumental SNPs were included. Decomposition in joint (βX, βY) effect space selected K = 3 components, yielding clear separation in two dimensions (Fig. 2B–C). Posterior assignments were highly concentrated for all but one SNP, indicating well-defined component boundaries.

**Fig. 2.**
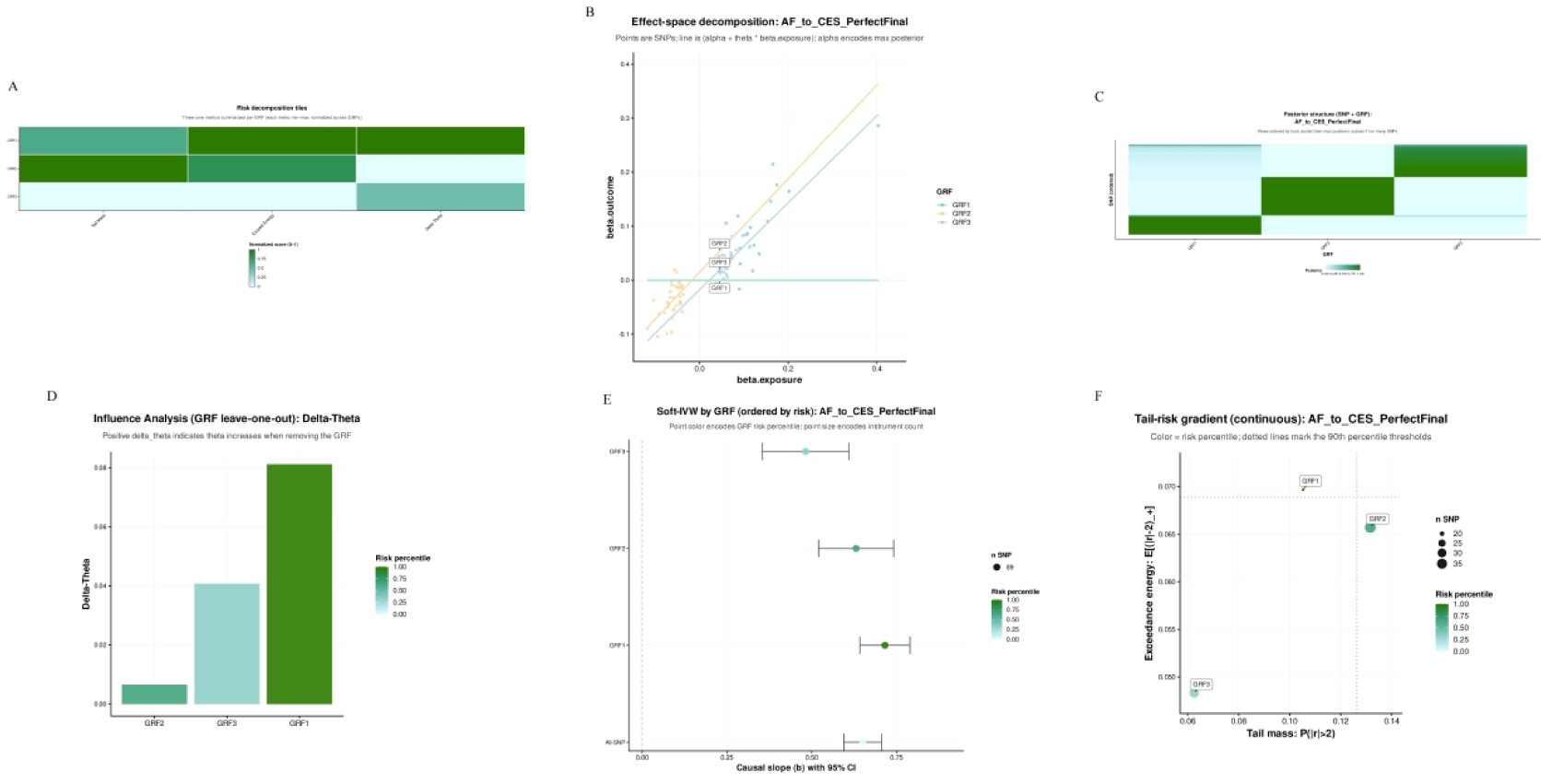
Effect-space decomposition and GRF-specific structure in AF → CES analysis. A, Risk decomposition across GRFs based on normalised tail-risk scores computed on |e|. B, Effect-space decomposition showing GRF-specific component structure in (βX, βY) space. C, Posterior assignment matrix of SNPs to GRFs. D, Component influence (GRF leave-one-out): change in the global slope θ after removing each GRF. E, GRF-specific soft-IVW estimates with 95% confidence intervals (ordered by slope magnitude). F, Tail-risk gradient across GRFs: tail mass versus exceedance energy.

At the effect level, GRF-specific soft-IVW estimates exhibited a graded amplitude hierarchy (Fig. 2E). GRF1 showed the largest slope (b = 0.716, 95% CI 0.642–0.790), followed by GRF2 (b = 0.631, 95% CI 0.520–0.742) and GRF3 (b = 0.482, 95% CI 0.354–0.609), establishing a consistent ordering (GRF1 > GRF2 > GRF3). All GRF estimates were obtained from the same SNP set under posterior weighting; differences across GRFs were expressed through the posterior weight distribution rather than input selection (hard memberships shown in Fig. 2C). Heterogeneity diagnostics indicated mild within-component variation in GRF1 and GRF2, with no evidence of systematic directional bias from intercept testing.

Tail-risk profiling further differentiated component structure (Fig. 2F). Tail-risk statistics were computed on the standardised residual magnitude |e|. GRF2 exhibited the highest tail mass, indicating a larger fraction of SNPs entering the extreme region, whereas GRF1 showed the highest exceedance energy, indicating more concentrated tail intensity. Tail-risk rankings were concordant with the amplitude hierarchy and component influence ordering, providing a distributional characterisation beyond slope magnitude alone.

Component-level leave-one-out analysis quantified structural influence directly (Fig. 2D). Removing GRF1 produced the largest reduction in the global slope (Δθ = 0.081), exceeding the shifts induced by removing GRF3 (Δθ = 0.041) or GRF2 (Δθ = 0.007). By contrast, the largest single-SNP leave-one-out shift was 0.020, substantially smaller than component-level shifts. Influence is component-driven rather than SNP-driven.

The risk decomposition matrix (Fig. 2A) summarises these hierarchical patterns across dimensions. Collectively, the AF→CES effect space exhibits a stable three-component structure with stratification in both effect magnitude and tail-risk architecture, rendering explicit structural heterogeneity not captured by a single global slope.

### 3.3 Systematic comparison of MR-UBRA with traditional MR and MR-Clust

In the AF→CES analysis, traditional MR using the default effective-tool filter in TwoSampleMR (mr_keep) included 87 SNPs and yielded consistent evidence for a positive overall association: IVW OR = 1.93 (1.81–2.06, P < 2e−16), MR-Egger OR = 2.18 (1.93–2.45, P < 2e−16), and weighted median OR = 2.03 (1.82–2.26, P < 2e−16) (Fig. 3A; Table 1). These estimators were concordant in direction and magnitude, but each represented the relationship with a single overall effect.

**Fig. 3.**
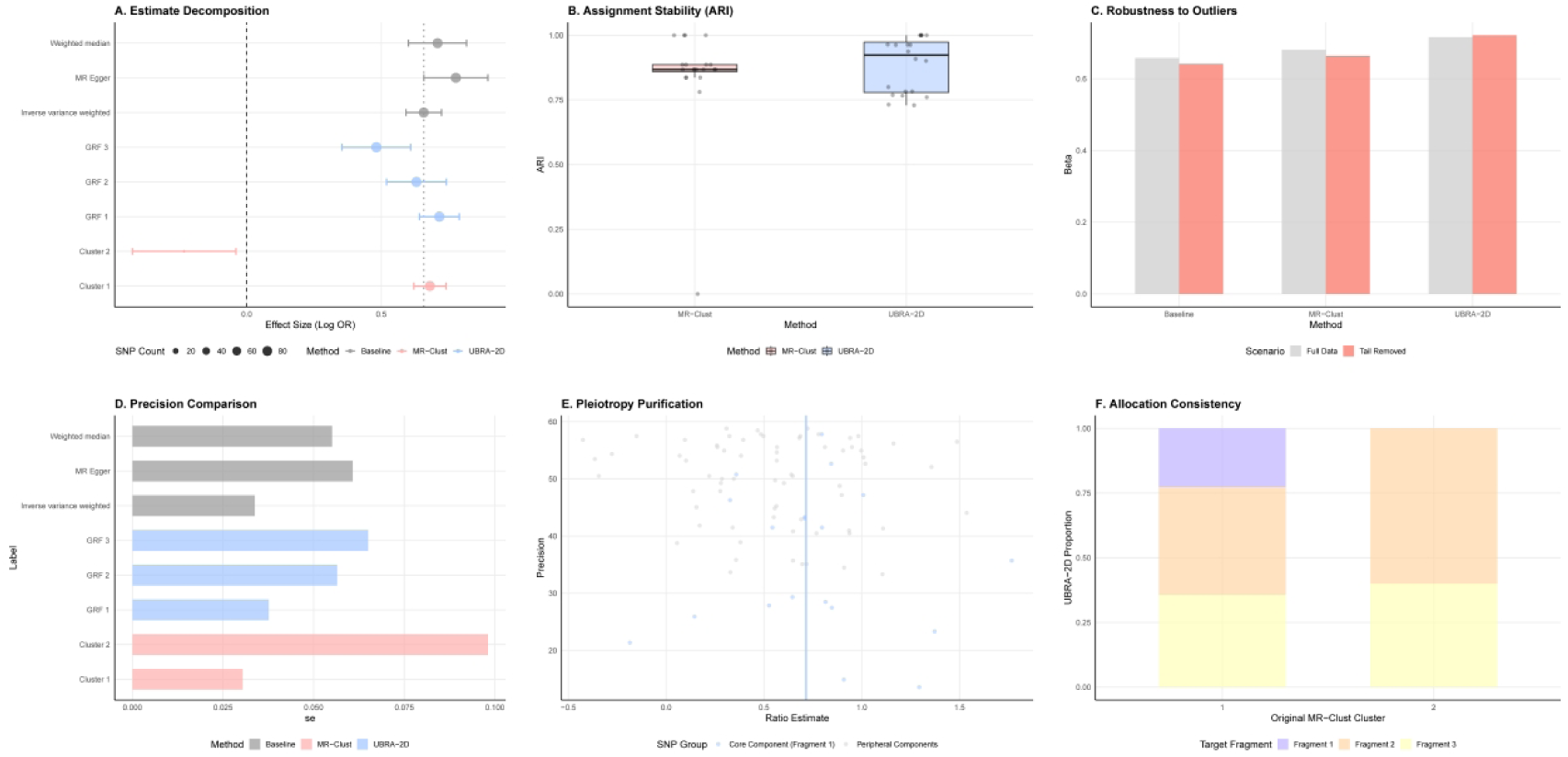
Method comparison and structural stability analyses. A, Causal effect estimates across traditional MR, MR-Clust, and MR-UBRA (effect sizes shown as log OR), including GRF-specific estimates. B, Assignment stability across repeated runs, quantified by the adjusted Rand index (ARI). C, Sensitivity to extreme effects: estimates under the full dataset and under tail-removed scenarios. D, Precision comparison across methods and components (standard errors). E, Ratio-estimate stratification for core versus peripheral SNP components. F, Cross-method allocation mapping between MR-Clust clusters and MR-UBRA fragments.

**Table 1.**
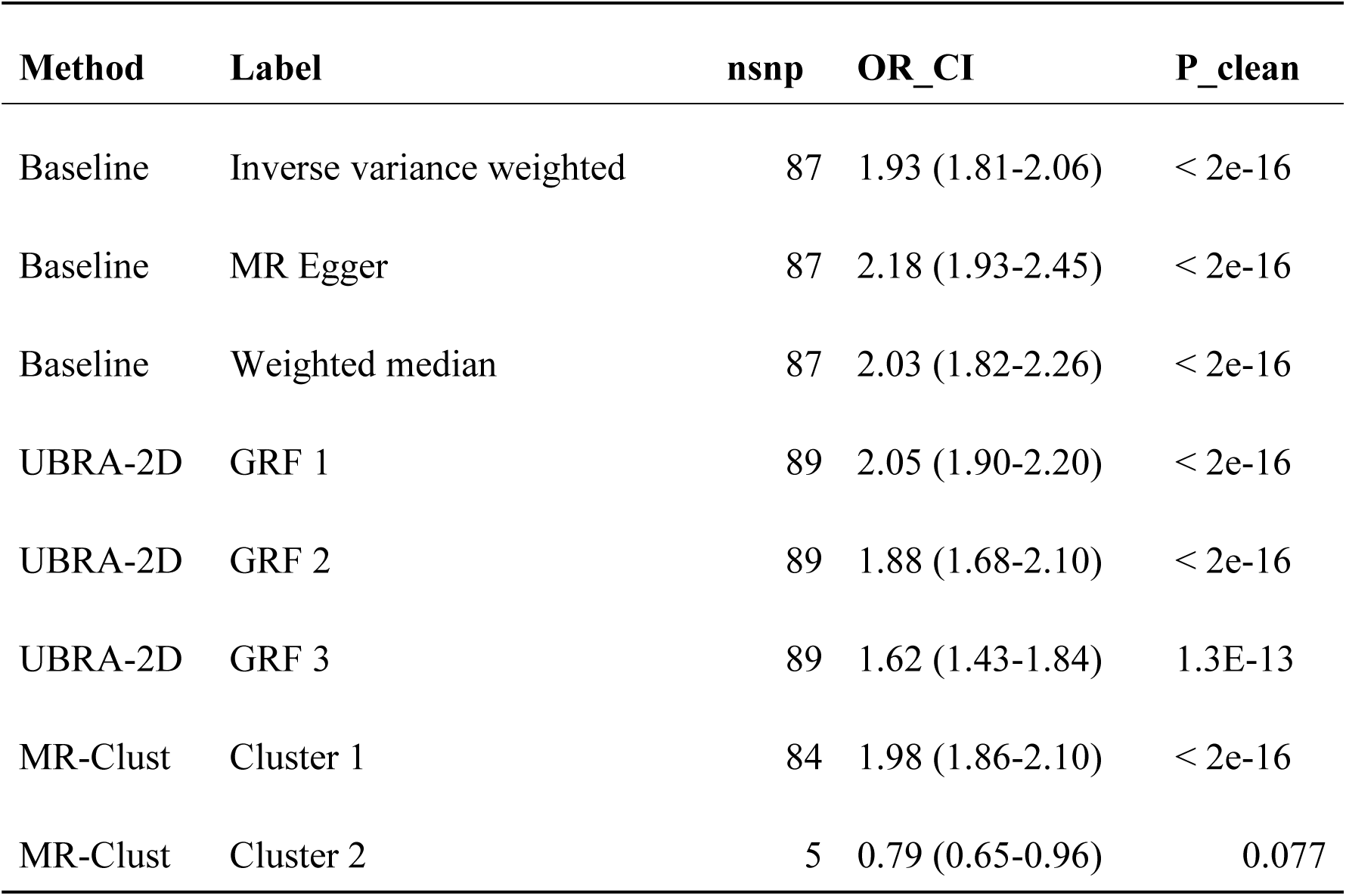
Comparative causal estimates across traditional MR, MR-Clust, and MR-UBRA in the AF→CES analysis. Odds ratios (ORs) with 95% confidence intervals (CIs) are reported. For MR-UBRA, GRF-specific effects are estimated using posterior-weighted soft-IVW based on the full harmonised SNP set.

On the same harmonised dataset (n = 89), MR-Clust identified two clusters: a dominant positive cluster (84 SNPs; OR = 1.98 (1.86–2.10), P < 2e−16) and a small secondary cluster (5 SNPs; OR = 0.79 (0.65–0.96), P = 0.077) (Fig. 3A; Table 1). In contrast, MR-UBRA (UBRA-2D) selected K = 3 and decomposed the positive association into a graded GRF-level amplitude hierarchy: GRF1 OR = 2.05 (1.90–2.20, P < 2e−16), GRF2 OR = 1.88 (1.68–2.10, P < 2e−16), and GRF3 OR = 1.62 (1.43–1.84, P = 1.3 × 10⁻¹³) (Fig. 3A; Table 1). MR-Clust partitions by direction; MR-UBRA partitions by structural amplitude.

For MR-UBRA, the nsnp value in Table 1 denotes the full harmonised SNP set used for posterior-weighted soft-IVW estimation; it therefore does not represent hard GRF membership counts and is not partitioned across GRFs as in MR-Clust (Fig. 3F; Tables 1–2).

**Table 2.** SNP accounting and weighting schemes across IVW, MR-Egger, MR-Clust and MR-UBRA in the AF→CES analysis.

| Method | SNPs<br>(Initial) | Post-clump | Post-QC | Used | Excluded | Weights |
| --- | --- | --- | --- | --- | --- | --- |
| IVW | 95 | 95 | 89 | 87 | TwoSampleMR<br>mr_keep filtering (2<br>SNPs) | Inverse<br>variance |
| MR-Egger | 95 | 95 | 89 | 87 | TwoSampleMR<br>mr_keep filtering (2<br>SNPs) | Inverse<br>variance |
| MR-Clust | 95 | 95 | 89 | 89 | None | Hard<br>clustering |
| MR-UBRA<br>(Proposed) | 95 | 95 | 89 | 89 | None | Soft<br>posterior |

Fig. 3B–F summarises assignment stability (ARI), sensitivity to tail/extreme-effect removal, precision (standard error) comparisons, separation in the precision–effect plane, and MR-Clust→MR-UBRA allocation mapping.

Note: Baseline traditional MR estimates (n = 87) follow the TwoSampleMR default mr_keep filter, whereas MR-Clust and MR-UBRA use the full harmonised SNP set (n = 89) for decomposition (Table 2), reflecting method-specific input definitions.

### 3.4 GRF stratification reveals multi-dimensional functional risk architecture in AF→CES

Following GRF decomposition of the AF→CES association, we assessed functional correspondence by performing SNP-to-gene mapping and pathway enrichment for each GRF under a unified framework (Fig. 4A–E). Enrichment profiles showed reproducible differences across GRFs and aligned with effect-space structure, including tail-risk characteristics and component-level influence on the global estimate, thereby providing a functional stratification of the average AF→CES association.

**Fig. 4.**
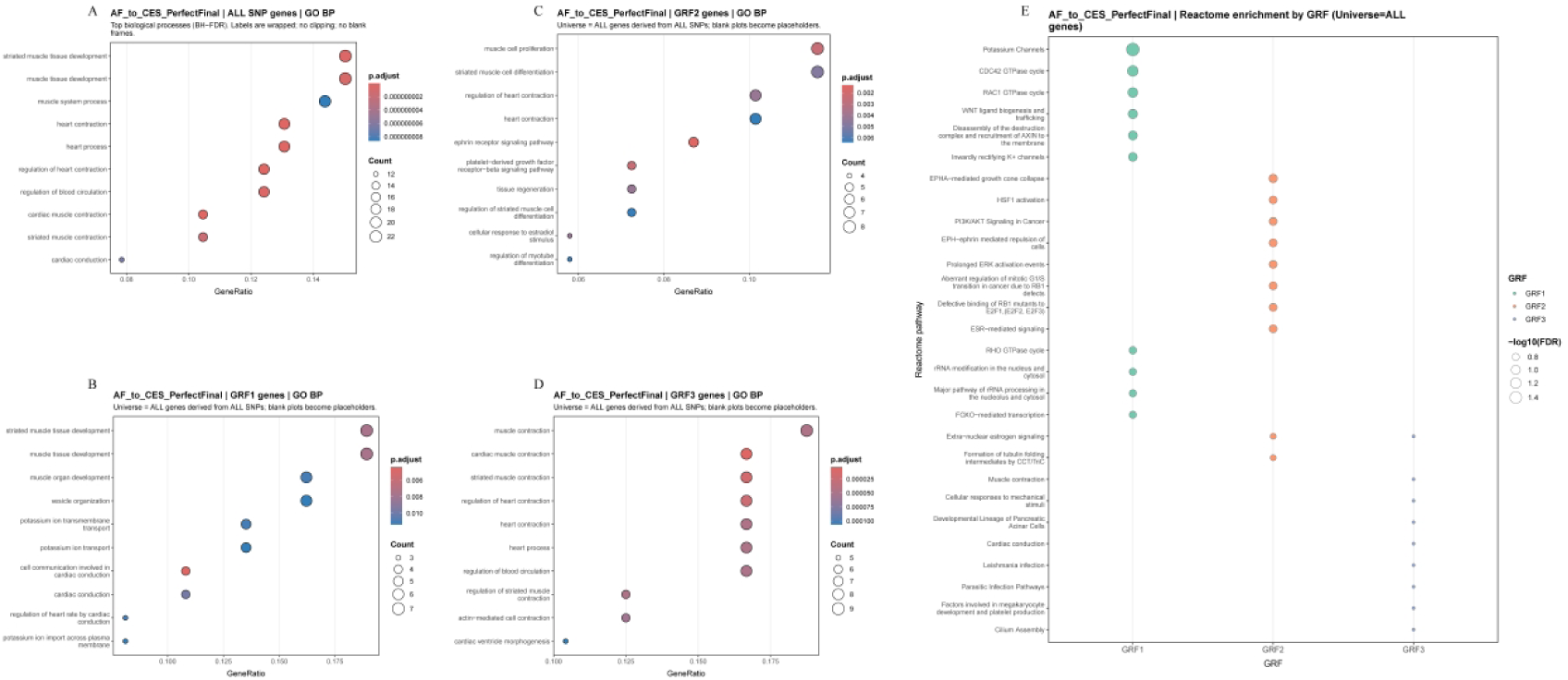
Functional enrichment across GRF-specific gene sets in AF→CES. A, Gene Ontology Biological Process (GO BP) enrichment for the full set of SNP-mapped genes (Benjamini–Hochberg FDR-adjusted). B, GO BP enrichment for the GRF1-specific SNP-mapped gene set. C, GO BP enrichment for the GRF2-specific SNP-mapped gene set. D, GO BP enrichment for the GRF3-specific SNP-mapped gene set. E, Reactome pathway enrichment across GRFs, using all SNP-mapped genes as the background set. Point size indicates -log10(FDR), and colour denotes GRF.

The high-impact core component (GRF1) was enriched for cardiac conduction and ion-channel related processes, including “cell communication involved in cardiac conduction” (36.3-fold enrichment, q = 0.003) and “potassium ion transmembrane transport” (11.8-fold, q = 0.008). Representative mapped genes included HCN4, KCNJ5, KCNJ1, and KCNN3. Consistent with Section 3.2, GRF1 exhibited the highest tail intensity and the largest component-level influence on the global estimate, in line with its conduction/channel-enriched annotation profile.

The intermediate-risk component (GRF2) was enriched for signalling and structural regulation pathways, including ephrin and PDGF receptor signalling (19.2-fold and 21.6-fold enrichment, q ≤ 0.002). Representative mapped genes included EPHA3, ERBB4, IGF1R, and MYOCD. Relative to GRF1, GRF2 contributed less to the global estimate and showed higher tail participation but lower tail intensity, corresponding to a broader signalling/structural annotation profile.

The low-impact component (GRF3) showed the smallest tail contribution and the lowest effect magnitude, with enrichment in contraction-related processes such as “cardiac muscle contraction” and “striated muscle contraction” (16–23-fold enrichment, q < 5 × 10⁻⁵). Representative mapped genes included MYH6, MYH7, and CASQ2, consistent with sarcomeric and calcium-handling modules. GRF3 corresponded to a contraction-related annotation theme with limited component-level influence on the global estimate.

Collectively, MR-UBRA stratifies the average AF→CES association into three hierarchically organised functional themes aligned with component structure: a conduction/channel-enriched component with the highest tail intensity and influence (GRF1), a signalling/structural component with broader tail participation (GRF2), and a contraction-enriched component with the lowest influence (GRF3). This statistical–functional correspondence links component-level structure to pathway-level annotations and supports component-aware downstream analyses.

### 3.5 Generalisation under strong heterogeneity: smoking → lung cancer

In the smoking → lung cancer association, traditional MR estimators provided concordant evidence for a positive overall association, while differing in effect magnitude and uncertainty (Table 3). IVW yielded OR = 1.58 (95% CI 1.28–1.94, P = 1.60 × 10⁻⁵) and the weighted median gave OR = 1.71 (1.30–2.26, P = 1.40 × 10⁻⁴), whereas MR-Egger returned a substantially wider confidence interval (OR = 1.46, 0.50–4.20, P = 0.490), reflecting higher estimator uncertainty. Together, these patterns are consistent with structured heterogeneity beyond a single-slope summary.

**Table 3.** Comparative causal estimates across traditional MR, MR-Clust, and MR-UBRA in the smoking → lung cancer analysis. Odds ratios (ORs) with 95% confidence intervals (CIs) are reported. For MR-UBRA, component-specific effects are estimated using posterior-weighted soft-IVW under the full harmonised SNP set.

| Method | Label | nsnp | OR_CI | P_clean |
| --- | --- | --- | --- | --- |
| Baseline | Inverse variance weighted | 70 | 1.58 (1.28-1.94) | 1.60E-05 |
| Baseline | MR Egger | 70 | 1.46 (0.50-4.20) | 0.48978 |
| Baseline | Weighted median | 70 | 1.71 (1.30-2.26) | 0.00014 |
| UBRA-2D | GRF 1 | 70 | 1.64 (1.28-2.10) | 8.10E-05 |
| UBRA-2D | GRF 2 | 70 | 1.51 (1.17-1.96) | 0.00169 |
| MR-Clust | Cluster 1 | 48 | 2.54 (2.17-2.97) | 2.30E-15 |
| MR-Clust | Cluster 2 | 22 | 0.60 (0.47-0.77) | 0.00051 |

Using the same harmonised input (Initial = 87; post-clumping = 87; used = 70; Table 4), MR-Clust partitioned instruments into two clusters with opposing directions (Table 3), yielding a directional split representation (Fig. 5A). In contrast, MR-UBRA identified two components in joint effect space (Table 3) that were directionally concordant yet stratified in amplitude, producing a graded “stronger–weaker” decomposition rather than a directional dichotomy (Fig. 5A).

**Fig. 5.**
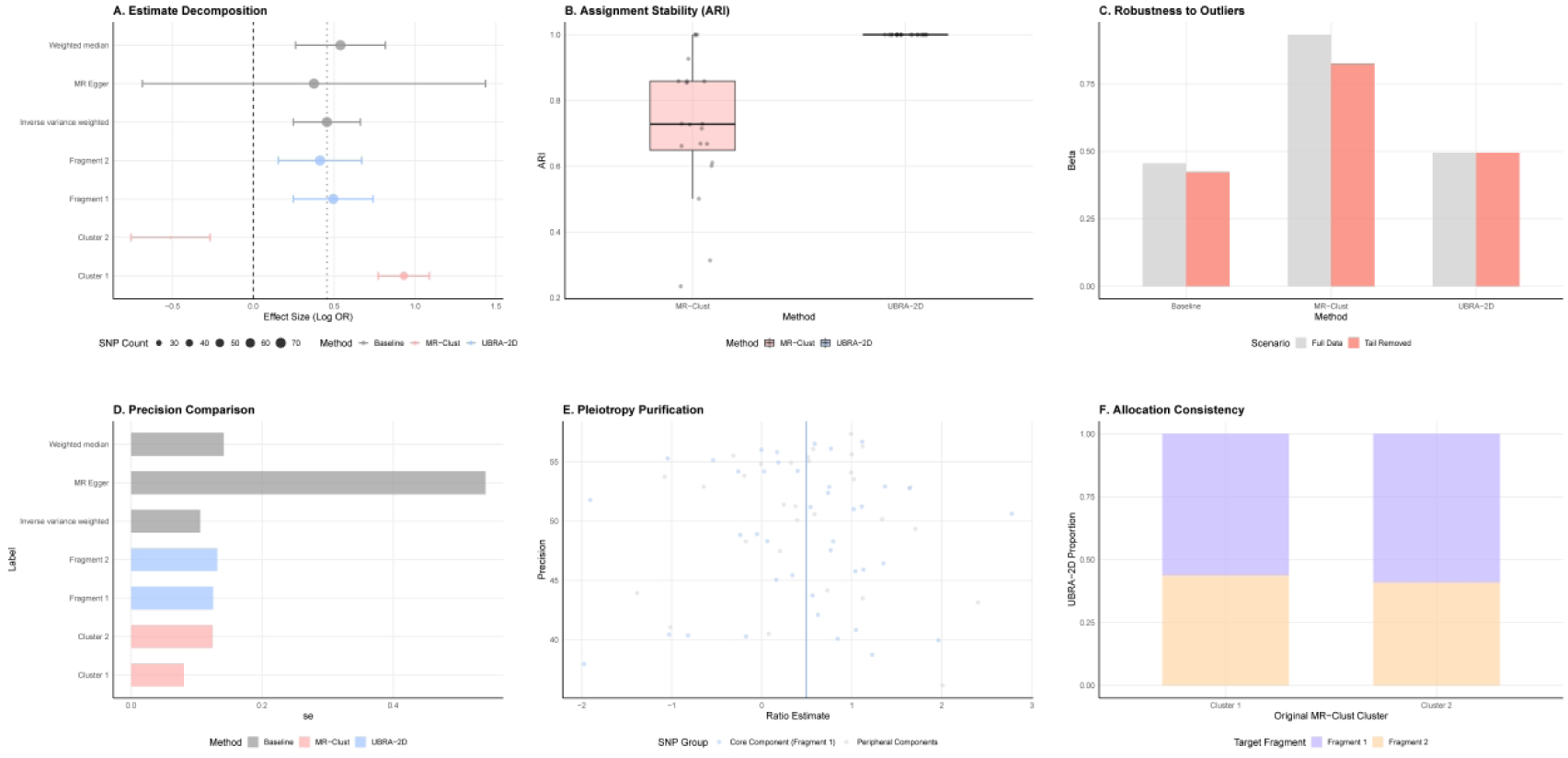
Performance under heterogeneous causal structure: smoking → lung cancer. A, Causal effect estimates across traditional MR, MR-Clust clusters, and MR-UBRA components (effect sizes shown as log OR). B, Assignment stability under resampling, quantified by the adjusted Rand index (ARI). C, Sensitivity to tail removal: estimates under the full dataset and under tail-removed scenarios. D, Precision comparison across estimators and components (standard errors). E, Ratio-estimate stratification for core versus peripheral SNP components. F, Cross-method allocation mapping between MR-Clust clusters and MR-UBRA components.

**Table 4.** SNP accounting and weighting schemes across IVW, MR-Egger, MR-Clust and MR-UBRA in the smoking→lung cancer analysis.

| Method | SNPs (Initial) | Post-clump | Used | Excluded | Weights |
| --- | --- | --- | --- | --- | --- |
| IVW | 87 | 87 | 70 | None | Inverse variance |
| MR-Egger | 87 | 87 | 70 | None | Inverse variance |
| MR-Clust | 87 | 87 | 70 | None | Hard clustering |
| MR-UBRA<br>(Proposed) | 87 | 87 | 70 | None | Soft posterior |

**Table 5.**
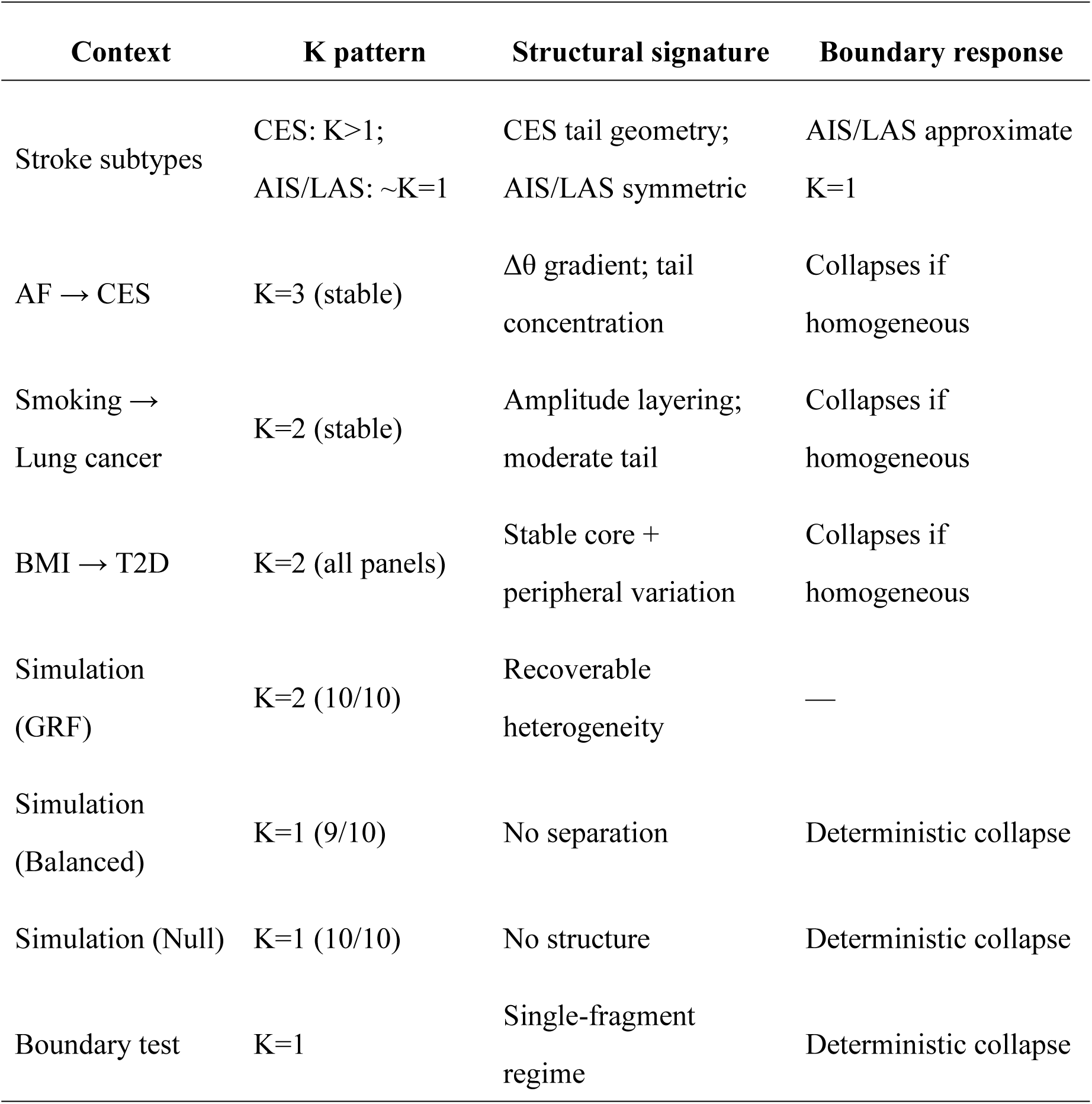
Cross-context structural behaviour and boundary consistency of MR-UBRA.

SNP accounting was identical across methods (Table 4), and differences therefore reflect aggregation schemes rather than input definition: IVW and MR-Egger use inverse-variance weighting, MR-Clust applies hard cluster assignment, and MR-UBRA uses posterior-weighted soft allocation for estimation.

Stability analyses further distinguished the two decomposition approaches (Fig. 5B–C). MR-UBRA exhibited near-deterministic assignment stability under resampling (ARI close to 1), whereas MR-Clust showed a broader ARI distribution. Under tail/extreme-effect removal perturbations, MR-UBRA estimates changed less than MR-Clust, indicating greater robustness to tail-focused perturbation in this heterogeneous setting.

Consistent with these empirical patterns, simulation boundary checks showed parsimonious behaviour across regimes: the model selected K = 1 under single-component structures, selected K = 2 under two-component structures, and reverted to K = 1 under extreme outlier pressure, maintaining a low false-positive split rate. Together, the smoking → lung cancer analysis supports a stable two-component representation and appropriate boundary behaviour of MR-UBRA under strong heterogeneity.

### 3.6 Consistent causal core across cohorts and ancestries: BMI → T2D analysis

Across European cohorts and cross-ancestry settings, BMI → T2D exhibited a consistently positive causal direction, whereas conventional IVW estimates in East Asian cohorts showed greater dispersion in effect magnitude (Fig. X A), indicating cohort- and instrument-dependent variability in single-slope summaries.

MR-UBRA partitioned this variability into a stable causal core (F1) and ancestry-sensitive tail components. Across European, East Asian, and cross-ancestry analyses, F1 retained a consistent direction with comparable effect size, while IVW estimates varied more across settings, indicating that between-cohort differences were primarily attributable to non-core structure (Fig. X A).

Tail-risk stratification further separated components by stability: increasing tail-risk energy (exceedance energy) was accompanied by amplified effect drift in tail components (e.g., F2 and other high-energy components), whereas the core component exhibited minimal drift across settings (Fig. X B).

In joint effect space (β_BMI, β_T2D), the core component maintained a consistent slope direction across ancestries, while tail components were more dispersed and exerted greater leverage on the overall estimate (Fig. X C).

Collectively, these results support a “stable core + ancestry-sensitive tail” architecture, making explicit the component structure underlying cross-cohort variation that is compressed in a single average slope.

### 3.7 Simulation-based evaluation of robustness and discriminative capacity under multi-mechanism settings

Under scenarios with coexisting mechanisms, the joint effect space (β_exposure, β_outcome) exhibited two linear structures with distinct slopes (Fig. 6A). The two-dimensional point cloud separated into two components occupying distinct regions of the joint distribution.

**Fig. 6.**
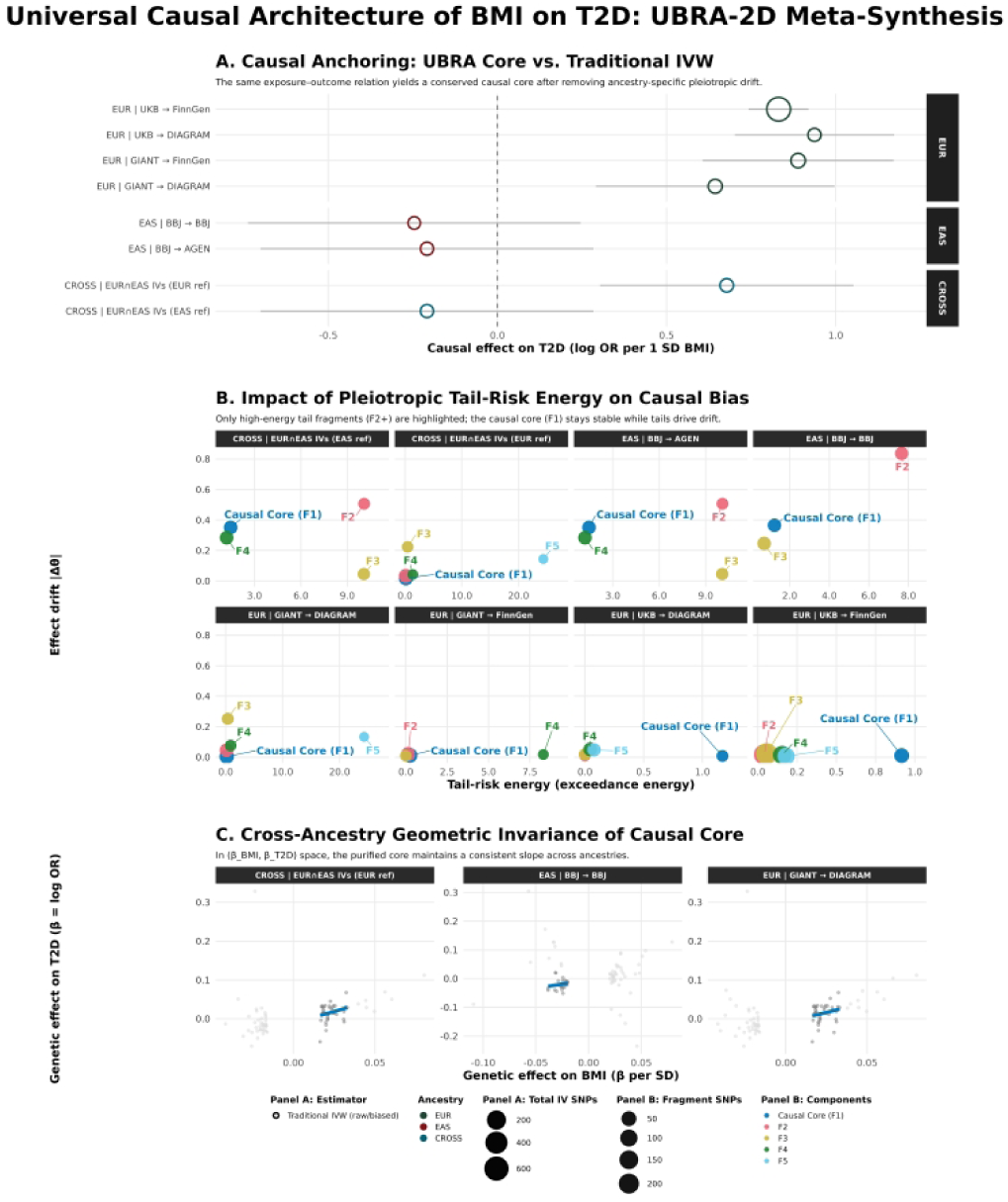
Cross-ancestry causal core in BMI → T2D. A, Causal effect estimates across European, East Asian, and cross-ancestry datasets, showing MR-UBRA core (F1) estimates alongside conventional IVW estimates (effect size shown as log OR per 1 SD increase in BMI). B, Association between tail-risk energy (exceedance energy) and effect drift across fragments: higher-energy fragments show greater drift, whereas the causal core (F1) exhibits minimal drift across datasets. C, Joint effect-space geometry (β_BMI, β_T2D) illustrating cross-ancestry consistency in the slope direction of the causal core (F1).

**Fig. 7.**
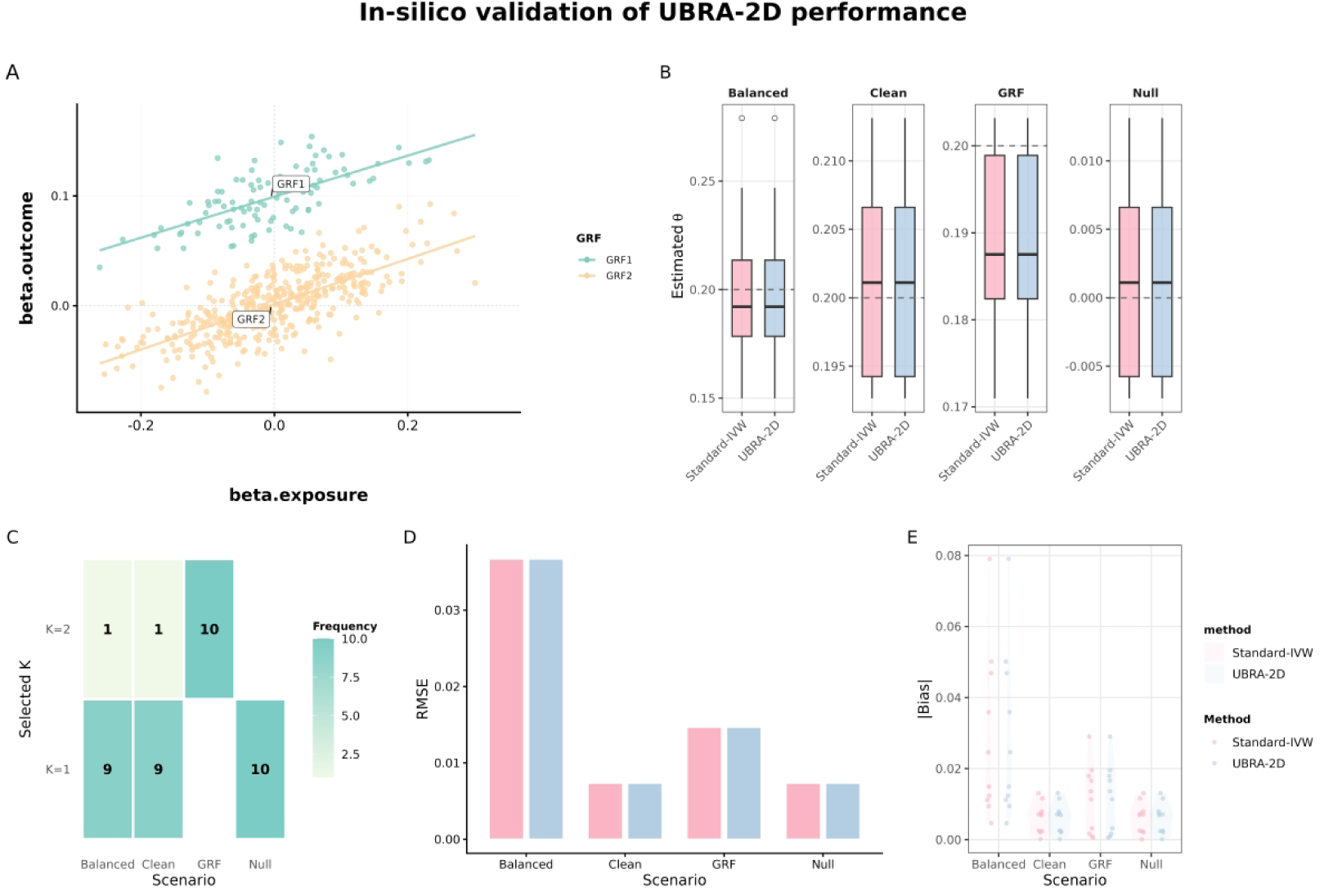
In-silico validation under multi-mechanism causal structures. A, Simulated joint effect space (β_exposure, β_outcome) illustrating two linear components under a multi-mechanism setting. B, Distribution of causal effect estimates across scenarios (Balanced, Clean, GRF, Null) for Standard-IVW and UBRA-2D. C, Model selection frequencies (K) across repeated simulations. D, Root mean squared error (RMSE) across scenarios. E, Bias distribution across scenarios.

At the estimation level (Fig. 6B, 6D–E), UBRA-2D and Standard-IVW yielded comparable estimate distributions across Balanced, Clean, and GRF settings. Across 10 simulation replicates:

Balanced (θ = 0.200): mean(θ^) = 0.201142; Bias = 0.001142; RMSE = 0.036694

Clean (θ = 0.200): mean(θ^) = 0.201371; Bias = 0.001371; RMSE = 0.007340

GRF (θ = 0.190): mean(θ^) = 0.189198; Bias = −0.000802; RMSE = 0.009945

Absolute biases were ≤ 0.001371 across structured settings.

Model selection results are shown in Fig. 6C. In the GRF scenario, UBRA-2D selected K = 2 in 10/10 replicates. In Balanced and Clean settings, K = 1 was selected in 9/10 replicates and K = 2 in 1/10. In the Null scenario, K = 1 was selected in 10/10 replicates. Standard-IVW corresponds to K = 1 in all scenarios.

Under the Null regime (θ = 0), both methods produced symmetric estimate distributions centred near zero (Fig. 6B, 6D–E), with mean(θ^) = 0.001371; Bias = 0.001371; RMSE = 0.007340.

Model complexity varied by structural configuration: multi-component settings yielded K = 2, whereas homogeneous and null architectures yielded K = 1. The global estimate corresponded to the posterior-weighted aggregation of component-specific effects.

### 3.8 Boundary behaviour under a single-component genetic architecture

To assess model behaviour under a single-component architecture, a boundary scenario containing only one causal component was constructed. Under this setting, the model consistently selected K = 1 according to both BIC and stability criteria (Fig. 8A), and effect-space decomposition yielded a single GRF. The point cloud in joint (β_exposure, β_outcome) space formed a continuous distribution without separable multi-component structure.

**Fig. 8.**
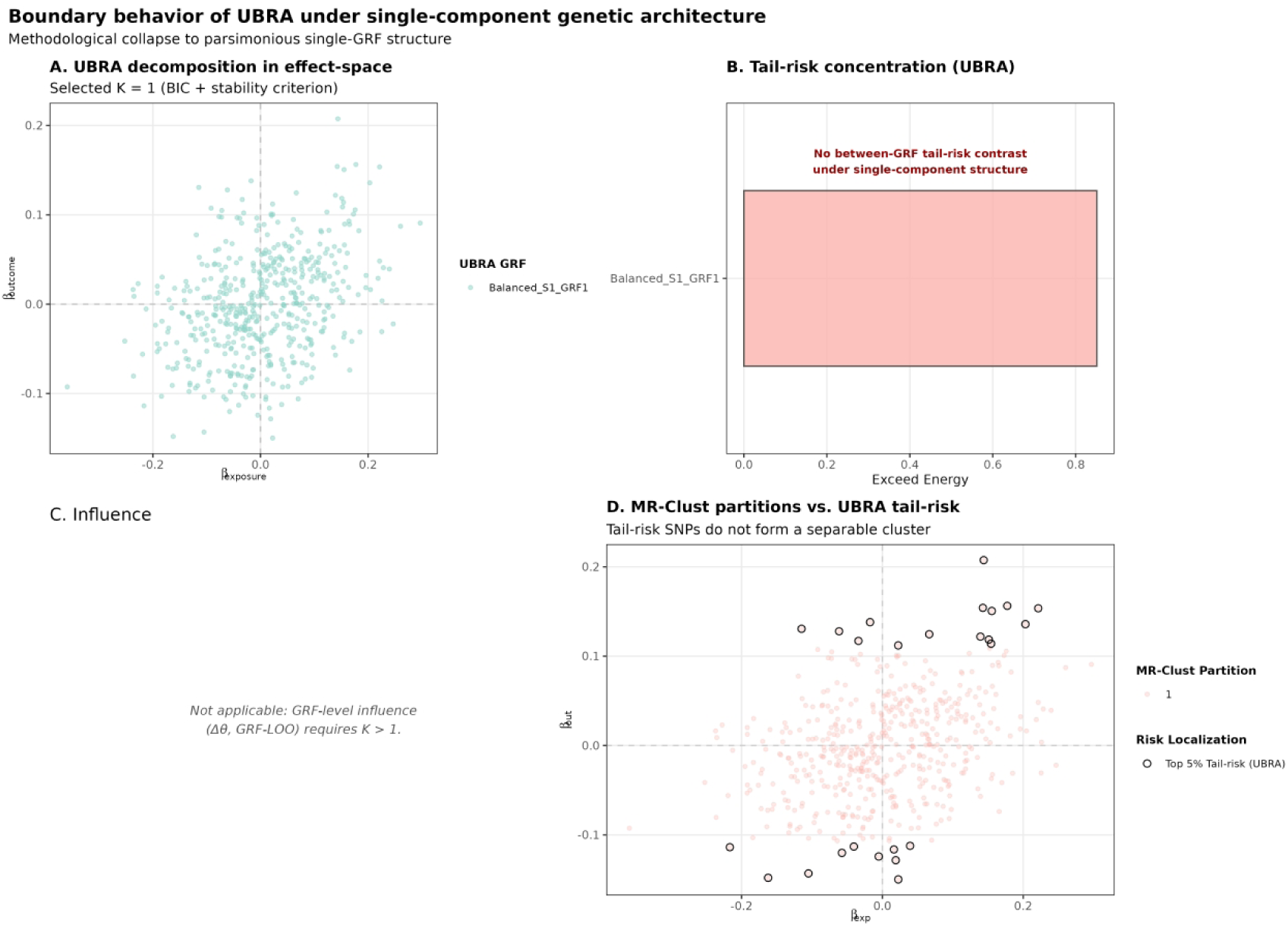
Boundary behaviour under single-component genetic architecture. A, Effect-space decomposition under a single-component structure (K = 1). B, Tail-risk concentration under K = 1 (no between-component contrast). C, GRF-level influence analysis (not applicable when K = 1). D, Distribution of top tail-risk SNPs in effect space, continuous rather than separable.

With only one GRF, no between-component tail-risk comparison was applicable (Fig. 8B). Tail-risk values were continuously distributed at the SNP level rather than concentrated within distinct structural units. Component-level influence analysis (Δθ, GRF-LOO) was not defined in this setting, as it requires K > 1 for decomposition (Fig. 8C).

High tail-risk SNPs (e.g., top 5%) were identifiable; however, these variants did not form separable clusters in effect space (Fig. 8D), but were embedded within the overall continuous distribution.

Model complexity remained at K = 1 throughout the boundary regime.

### 3.9 Consistent structural behaviour across empirical and simulated settings

Across empirical and simulated analyses, model complexity varied according to the degree of heterogeneity in joint effect space (Table X). In settings with structural heterogeneity, multi-component representations were selected under resampling and perturbation. Under homogeneous or null conditions, the model converged to a single-component solution (K = 1).

In stroke subtype analyses, CES exhibited multi-component structure with concentrated tail energy, whereas AIS and LAS showed near-symmetric distributions approximating a single-component structure. In AF→CES and smoking→lung cancer, multi-component decompositions were observed, characterised by amplitude gradients and concentrated tail energy. Across eight cross-ancestry BMI→T2D configurations, a two-component structure was observed, comprising a core component reproducible across datasets and a peripheral component exhibiting ancestry-sensitive variation.

In simulation analyses, multi-component structures were selected in GRF scenarios with heterogeneous architecture. Under balanced or null scenarios, the model converged to K = 1 in the majority of replicates.

Across empirical, cross-ancestry, and simulated settings, component number corresponded to structural configuration in joint effect space.

## 4 Discussion

### 4.1 Effect-space representation of causal structure

Causal effects in Mendelian randomization are commonly summarised as a single projection slope describing directional association between exposure and outcome[43–45]. This representation is well suited to homogeneous and symmetric genetic architectures in joint (βX, βY) space[46]. Under heterogeneous architectures, effect magnitudes may exhibit stratified or asymmetric organisation rather than diffuse dispersion around a central trend[47, 48].

Across empirical settings, effect-space geometry displayed component stratification and amplitude layering[48]. Projection-based summaries compress this geometry into a scalar quantity[43]. Representing causal effects as structured distributions in joint effect space introduces an additional descriptive layer while preserving the classical MR estimand[43, 49].

In this formulation, heterogeneity is expressed as spatial organisation within effect space, and single-slope estimation constitutes one projection of a potentially multi-component structure[50, 51].

### 4.2 Heterogeneity as structured signal

Within conventional MR analyses, heterogeneity is typically addressed as residual variation[50]. In the present framework, heterogeneity is represented in joint effect space as reproducible component clustering and hierarchical differentiation[45, 48].

When effect magnitudes form separable clusters, variation is organised rather than diffuse[47, 48]. Decomposition in effect space partitions this organisation into components without modifying directional inference or the global estimand[51, 52].

Under homogeneous architectures, such spatial organisation is not observed and the model converges to a single component. Under heterogeneous architectures, component differentiation reflects structured variation in effect magnitude and distribution. Heterogeneity is thus represented as geometry rather than residual dispersion[50].

### 4.3 Tail dominance as a distributional property

In several trait contexts, the distribution of standardised residual magnitude |e| exhibited skewness or heavy-tailed structure[46, 53]. Variants occupying extreme regions of effect space contributed disproportionately to distributional geometry[47, 48].

Tail dominance is defined relative to the backbone projection and characterised by concentration of residual magnitude within specific regions of effect space[54]. This property reflects collective spatial concentration rather than isolated outliers. Tail metrics quantify this concentration without altering slope estimation.

Slope estimates summarise directional association; tail metrics describe deviation structure. These descriptors operate at distinct representational levels within the same causal framework.

### 4.4 Structural–functional alignment

In selected analyses, component stratification in effect space corresponded to distinct functional enrichment profiles[55–57]. Statistical partitioning aligned with pathway-level annotations under the same decomposition[55, 58].

Such correspondence reflects concordance between spatial organisation and annotation themes. It does not constitute mechanistic inference, as enrichment depends on existing annotation systems[59, 60]. Structural stratification and functional annotation therefore remain descriptive layers operating in parallel.

Effect-space decomposition provides a statistical partition that can be examined alongside functional information without implying component-level mechanism[56, 61].

### 4.5 Structural behaviour across ancestry contexts

In cross-ancestry analyses, effect-space decomposition separated a reproducible core component from peripheral components exhibiting ancestry-sensitive variation[62–64]. Single-slope estimates aggregate contributions across these layers.

Differences in overall slope magnitude across cohorts can be expressed as variation in component weighting within joint effect space[63, 65, 66]. Structural decomposition describes this redistribution without redefining causal direction or altering the global estimand[64].

Causal effects are thus represented as layered spatial structures whose relative weights may vary across populations[67–69].

### 4.6 Adaptive behaviour under homogeneous architectures

Boundary analyses showed that under single-component genetic architecture, the model selected K = 1 and no additional structure emerged[53]. Effect-space decomposition yielded a single GRF and tail metrics were continuously distributed at the SNP level.

Model complexity corresponded to the presence or absence of spatial heterogeneity in joint effect space[50, 70]. Under homogeneous conditions, representation remained parsimonious; under heterogeneous conditions, component differentiation was observed.

Decomposition therefore functions as a conditional descriptive layer rather than an imposed partition.

### 4.7 Limitations and outlook

Identification of component structure depends on instrument number and effect distribution. Sparse instruments may limit resolution of effect-space stratification. Selection of component number relies on statistical criteria and stability diagnostics, and alternative criteria may yield different partitions in marginal cases[49, 52].

Functional enrichment analyses depend on current annotation resources and provide descriptive correspondence rather than mechanistic validation[56, 59]. Component stratification should therefore be interpreted at the level of spatial organisation in effect space.

Future work may evaluate reproducibility of effect-space organisation across broader trait domains and ancestry contexts, and explore higher-dimensional or dynamic effect representations[45, 62]. Such extensions would further situate causal effects within a geometric framework of structured variation while maintaining compatibility with the classical MR estimand.

## 5 Conclusion

This study presents MR-UBRA as a representation of causal effects in joint effect space while preserving the classical Mendelian randomization estimand[44]. By decomposing instrumental variants in (βX, βY) space through probabilistic mixture modelling, causal effects are expressed as structured distributions composed of genetic risk fragments rather than solely as projection slopes[47, 48].

Across empirical and simulated settings, effect-space decomposition identified component stratification under heterogeneous architectures and converged to single-component representation under homogeneous regimes[46, 51]. Structural organisation in joint effect space was compatible with directional slope inference and remained aligned with the original causal objective.

MR-UBRA therefore situates causal effects within a geometric framework of spatial organisation, in which heterogeneity is represented as structured variation rather than residual dispersion[49]. This representation extends descriptive resolution in complex genetic architectures while maintaining compatibility with conventional MR inference.

## Author Contributions

H. Hongdong conceived the study and developed the MR-UBRA framework.

H. Hongdong and D.Chen designed the statistical methodology and performed the primary analyses.

C. Qian, X.Zhou and H.Huang contributed to data curation, harmonisation and quality control.

J. Zuo, G.Wang and X.Peng assisted with simulation design, validation analyses and interpretation of the results.

H.-X. Liu supervised the project and provided conceptual guidance.

H.Hongdong drafted the manuscript. All authors reviewed, edited and approved the final version of the manuscript.

## Abbreviations

MR: Mendelian randomization
MR-UBRA: Mendelian randomization–Unified Bayesian Risk Architecture
GRF: genetic risk fragment
IVW: inverse variance weighted
VaR: value at risk
CVaR: conditional value at risk
CES: cardioembolic stroke
AIS: acute ischemic stroke
LAS: large artery stroke
BMI: body mass index
T2D: type 2 diabetes.

## Data Availability

All GWAS summary statistics used in this study are publicly available from the original consortium repositories cited in the manuscript. The MR-UBRA implementation and analysis code are available on GitHub at: https://github.com/0808hao/MR-UBRA. All derived data generated during this study are included in the manuscript and its supplementary materials.

https://www.strokegenetics.org/projects/megastroke/

https://giant-consortium.web.broadinstitute.org/GIANT_consortium_data_files

https://pheweb.jp/downloads

## References

1. Burgess S, Butterworth A, Thompson SG: Mendelian randomization analysis with multiple genetic variants using summarized data. Genet Epidemiol 2013, 37(7):658–665.

2. Davey Smith G, Hemani G: Mendelian randomization: genetic anchors for causal inference in epidemiological studies. Hum Mol Genet 2014, 23(R1):R89–98.

3. Burgess S, Scott RA, Timpson NJ, Davey Smith G, Thompson SG: Using published data in Mendelian randomization: a blueprint for efficient identification of causal risk factors. Eur J Epidemiol 2015, 30(7):543–552.

4. Pierce BL, Burgess S: Efficient design for Mendelian randomization studies: subsample and 2-sample instrumental variable estimators. Am J Epidemiol 2013, 178(7):1177–1184.

5. Qi G, Chatterjee N: Mendelian randomization analysis using mixture models for robust and efficient estimation of causal effects. Nat Commun 2019, 10(1):1941.

6. Verbanck M, Chen CY, Neale B, Do R: Detection of widespread horizontal pleiotropy in causal relationships inferred from Mendelian randomization between complex traits and diseases. Nat Genet 2018, 50(5):693–698.

7. Burgess S, Bowden J, Fall T, Ingelsson E, Thompson SG: Sensitivity Analyses for Robust Causal Inference from Mendelian Randomization Analyses with Multiple Genetic Variants. Epidemiology 2017, 28(1):30–42.

8. Bowden J, Davey Smith G, Burgess S: Mendelian randomization with invalid instruments: effect estimation and bias detection through Egger regression. Int J Epidemiol 2015, 44(2):512–525.

9. Foley CN, Mason AM, Kirk PDW, Burgess S: MR-Clust: clustering of genetic variants in Mendelian randomization with similar causal estimates. Bioinformatics 2021, 37(4):531–541.

10. Bowden J, Davey Smith G, Haycock PC, Burgess S: Consistent Estimation in Mendelian Randomization with Some Invalid Instruments Using a Weighted Median Estimator. Genet Epidemiol 2016, 40(4):304–314.

11. Solovieff N, Cotsapas C, Lee PH, Purcell SM, Smoller JW: Pleiotropy in complex traits: challenges and strategies. Nat Rev Genet 2013, 14(7):483–495.

12. Sivakumaran S, Agakov F, Theodoratou E, Prendergast JG, Zgaga L, Manolio T, Rudan I, McKeigue P, Wilson JF, Campbell H: Abundant pleiotropy in human complex diseases and traits. Am J Hum Genet 2011, 89(5):607–618.

13. O’Connor LJ, Price AL: Distinguishing genetic correlation from causation across 52 diseases and complex traits. Nat Genet 2018, 50(12):1728–1734.

14. Zhang Y, Qi G, Park JH, Chatterjee N: Estimation of complex effect-size distributions using summary-level statistics from genome-wide association studies across 32 complex traits. Nat Genet 2018, 50(9):1318–1326.

15. Park JH, Wacholder S, Gail MH, Peters U, Jacobs KB, Chanock SJ, Chatterjee N: Estimation of effect size distribution from genome-wide association studies and implications for future discoveries. Nat Genet 2010, 42(7):570–575.

16. Holland D, Frei O, Desikan R, Fan CC, Shadrin AA, Smeland OB, Sundar VS, Thompson P, Andreassen OA, Dale AM: Beyond SNP heritability: Polygenicity and discoverability of phenotypes estimated with a univariate Gaussian mixture model. PLoS Genet 2020, 16(5):e1008612.

17. Frei O, Holland D, Smeland OB, Shadrin AA, Fan CC, Maeland S, O’Connell KS, Wang Y, Djurovic S, Thompson WK et al: Bivariate causal mixture model quantifies polygenic overlap between complex traits beyond genetic correlation. Nat Commun 2019, 10(1):2417.

18. Boyle EA, Li YI, Pritchard JK: An Expanded View of Complex Traits: From Polygenic to Omnigenic. Cell 2017, 169(7):1177–1186.

19. Bulik-Sullivan B, Finucane HK, Anttila V, Gusev A, Day FR, Loh PR, Duncan L, Perry JR, Patterson N, Robinson EB et al: An atlas of genetic correlations across human diseases and traits. Nat Genet 2015, 47(11):1236–1241.

20. Cho Y, Haycock PC, Sanderson E, Gaunt TR, Zheng J, Morris AP, Davey Smith G, Hemani G: Exploiting horizontal pleiotropy to search for causal pathways within a Mendelian randomization framework. Nat Commun 2020, 11(1):1010.

21. Bowden J, Del Greco MF, Minelli C, Zhao Q, Lawlor DA, Sheehan NA, Thompson J, Davey Smith G: Improving the accuracy of two-sample summary-data Mendelian randomization: moving beyond the NOME assumption. Int J Epidemiol 2019, 48(3):728–742.

22. Spiller W, Bowden J, Sanderson E: Estimating and visualising multivariable Mendelian randomization analyses within a radial framework. PLoS Genet 2024, 20(12):e1011506.

23. Morrison J, Knoblauch N, Marcus JH, Stephens M, He X: Mendelian randomization accounting for correlated and uncorrelated pleiotropic effects using genome-wide summary statistics. Nat Genet 2020, 52(7):740–747.

24. Grant AJ, Burgess S: A Bayesian approach to Mendelian randomization using summary statistics in the univariable and multivariable settings with correlated pleiotropy. Am J Hum Genet 2024, 111(1):165–180.

25. Zuber V, Colijn JM, Klaver C, Burgess S: Selecting likely causal risk factors from high-throughput experiments using multivariable Mendelian randomization. Nat Commun 2020, 11(1):29.

26. Burgess S, Foley CN, Allara E, Staley JR, Howson JMM: A robust and efficient method for Mendelian randomization with hundreds of genetic variants. Nat Commun 2020, 11(1):376.

27. Sanderson E, Glymour MM, Holmes MV, Kang H, Morrison J, Munafò MR, Palmer T, Schooling CM, Wallace C, Zhao Q et al: Mendelian randomization. Nat Rev Methods Primers 2022, 2.

28. Bowden J, Spiller W, Del Greco MF, Sheehan N, Thompson J, Minelli C, Davey Smith G: Improving the visualization, interpretation and analysis of two-sample summary data Mendelian randomization via the Radial plot and Radial regression. Int J Epidemiol 2018, 47(4):1264–1278.

29. Burgess S, Thompson SG: Avoiding bias from weak instruments in Mendelian randomization studies. Int J Epidemiol 2011, 40(3):755–764.

30. McNicholas PD, Murphy TB: Model-based clustering of microarray expression data via latent Gaussian mixture models. Bioinformatics 2010, 26(21):2705–2712.

31. Vrieze SI: Model selection and psychological theory: a discussion of the differences between the Akaike information criterion (AIC) and the Bayesian information criterion (BIC). Psychol Methods 2012, 17(2):228–243.

32. Wilkerson MD, Hayes DN: ConsensusClusterPlus: a class discovery tool with confidence assessments and item tracking. Bioinformatics 2010, 26(12):1572–1573.

33. Steinley D: Properties of the Hubert-Arabie adjusted Rand index. Psychol Methods 2004, 9(3):386–396.

34. Müller FM, Righi MB: Comparison of Value at Risk (VaR) Multivariate Forecast Models. Comput Econ 2022:1–36.

35. Boca SM, Leek JT: A direct approach to estimating false discovery rates conditional on covariates. PeerJ 2018, 6:e6035.

36. Bowden J, Del Greco MF, Minelli C, Davey Smith G, Sheehan NA, Thompson JR: Assessing the suitability of summary data for two-sample Mendelian randomization analyses using MR-Egger regression: the role of the I2 statistic. Int J Epidemiol 2016, 45(6):1961–1974.

37. Higgins JP, Thompson SG, Deeks JJ, Altman DG: Measuring inconsistency in meta-analyses. Bmj 2003, 327(7414):557–560.

38. Malik R, Chauhan G, Traylor M, Sargurupremraj M, Okada Y, Mishra A, Rutten-Jacobs L, Giese AK, van der Laan SW, Gretarsdottir S et al: Multiancestry genome-wide association study of 520,000 subjects identifies 32 loci associated with stroke and stroke subtypes. Nat Genet 2018, 50(4):524–537.

39. Fadista J, Manning AK, Florez JC, Groop L: The (in)famous GWAS P-value threshold revisited and updated for low-frequency variants. Eur J Hum Genet 2016, 24(8):1202–1205.

40. Purcell S, Neale B, Todd-Brown K, Thomas L, Ferreira MA, Bender D, Maller J, Sklar P, de Bakker PI, Daly MJ et al: PLINK: a tool set for whole-genome association and population-based linkage analyses. Am J Hum Genet 2007, 81(3):559–575.

41. Hemani G, Tilling K, Davey Smith G: Orienting the causal relationship between imprecisely measured traits using GWAS summary data. PLoS Genet 2017, 13(11):e1007081.

42. Peng RD: Reproducible research in computational science. Science 2011, 334(6060):1226–1227.

43. Davies NM, Holmes MV, Davey Smith G: Reading Mendelian randomisation studies: a guide, glossary, and checklist for clinicians. Bmj 2018, 362:k601.

44. Smith GD, Ebrahim S: ‘Mendelian randomization’: can genetic epidemiology contribute to understanding environmental determinants of disease? Int J Epidemiol 2003, 32(1):1–22.

45. Skrivankova VW, Richmond RC, Woolf BAR, Yarmolinsky J, Davies NM, Swanson SA, VanderWeele TJ, Higgins JPT, Timpson NJ, Dimou N et al: Strengthening the Reporting of Observational Studies in Epidemiology Using Mendelian Randomization: The STROBE-MR Statement. Jama 2021, 326(16):1614–1621.

46. Visscher PM, Wray NR, Zhang Q, Sklar P, McCarthy MI, Brown MA, Yang J: 10 Years of GWAS Discovery: Biology, Function, and Translation. Am J Hum Genet 2017, 101(1):5–22.

47. Gratten J, Visscher PM: Genetic pleiotropy in complex traits and diseases: implications for genomic medicine. Genome Med 2016, 8(1):78.

48. Zhang Z, Jung J, Kim A, Suboc N, Gazal S, Mancuso N: A scalable approach to characterize pleiotropy across thousands of human diseases and complex traits using GWAS summary statistics. Am J Hum Genet 2023, 110(11):1863–1874.

49. Skrivankova VW, Richmond RC, Woolf BAR, Davies NM, Swanson SA, VanderWeele TJ, Timpson NJ, Higgins JPT, Dimou N, Langenberg C et al: Strengthening the reporting of observational studies in epidemiology using mendelian randomisation (STROBE-MR): explanation and elaboration. Bmj 2021, 375:n2233.

50. Hemani G, Bowden J, Davey Smith G: Evaluating the potential role of pleiotropy in Mendelian randomization studies. Hum Mol Genet 2018, 27(R2):R195–r208.

51. Cheng Q, Zhang X, Chen LS, Liu J: Mendelian randomization accounting for complex correlated horizontal pleiotropy while elucidating shared genetic etiology. Nat Commun 2022, 13(1):6490.

52. Bowden J, Del Greco MF, Minelli C, Davey Smith G, Sheehan N, Thompson J: A framework for the investigation of pleiotropy in two-sample summary data Mendelian randomization. Stat Med 2017, 36(11):1783–1802.

53. Schoech AP, Jordan DM, Loh PR, Gazal S, O’Connor LJ, Balick DJ, Palamara PF, Finucane HK, Sunyaev SR, Price AL: Quantification of frequency-dependent genetic architectures in 25 UK Biobank traits reveals action of negative selection. Nat Commun 2019, 10(1):790.

54. Lloyd-Jones LR, Zeng J, Sidorenko J, Yengo L, Moser G, Kemper KE, Wang H, Zheng Z, Magi R, Esko T et al: Improved polygenic prediction by Bayesian multiple regression on summary statistics. Nat Commun 2019, 10(1):5086.

55. de Leeuw CA, Mooij JM, Heskes T, Posthuma D: MAGMA: generalized gene-set analysis of GWAS data. PLoS Comput Biol 2015, 11(4):e1004219.

56. Watanabe K, Taskesen E, van Bochoven A, Posthuma D: Functional mapping and annotation of genetic associations with FUMA. Nat Commun 2017, 8(1):1826.

57. Pers TH, Karjalainen JM, Chan Y, Westra HJ, Wood AR, Yang J, Lui JC, Vedantam S, Gustafsson S, Esko T et al: Biological interpretation of genome-wide association studies using predicted gene functions. Nat Commun 2015, 6:5890.

58. Iotchkova V, Ritchie GRS, Geihs M, Morganella S, Min JL, Walter K, Timpson NJ, Dunham I, Birney E, Soranzo N: GARFIELD classifies disease-relevant genomic features through integration of functional annotations with association signals. Nat Genet 2019, 51(2):343–353.

59. Kundaje A, Meuleman W, Ernst J, Bilenky M, Yen A, Heravi-Moussavi A, Kheradpour P, Zhang Z, Wang J, Ziller MJ et al: Integrative analysis of 111 reference human epigenomes. Nature 2015, 518(7539):317–330.

60. Boyle AP, Hong EL, Hariharan M, Cheng Y, Schaub MA, Kasowski M, Karczewski KJ, Park J, Hitz BC, Weng S et al: Annotation of functional variation in personal genomes using RegulomeDB. Genome Res 2012, 22(9):1790–1797.

61. Finucane HK, Bulik-Sullivan B, Gusev A, Trynka G, Reshef Y, Loh PR, Anttila V, Xu H, Zang C, Farh K et al: Partitioning heritability by functional annotation using genome-wide association summary statistics. Nat Genet 2015, 47(11):1228–1235.

62. Li YR, Keating BJ: Trans-ethnic genome-wide association studies: advantages and challenges of mapping in diverse populations. Genome Med 2014, 6(10):91.

63. Mägi R, Horikoshi M, Sofer T, Mahajan A, Kitajima H, Franceschini N, McCarthy MI, Morris AP: Trans-ethnic meta-regression of genome-wide association studies accounting for ancestry increases power for discovery and improves fine-mapping resolution. Hum Mol Genet 2017, 26(18):3639–3650.

64. Brown BC, Ye CJ, Price AL, Zaitlen N: Transethnic Genetic-Correlation Estimates from Summary Statistics. Am J Hum Genet 2016, 99(1):76–88.

65. Brumpton B, Sanderson E, Heilbron K, Hartwig FP, Harrison S, Vie G, Cho Y, Howe LD, Hughes A, Boomsma DI et al: Avoiding dynastic, assortative mating, and population stratification biases in Mendelian randomization through within-family analyses. Nat Commun 2020, 11(1):3519.

66. Ma Y, Zhou X: Genetic prediction of complex traits with polygenic scores: a statistical review. Trends Genet 2021, 37(11):995–1011.

67. Popejoy AB, Fullerton SM: Genomics is failing on diversity. Nature 2016, 538(7624):161–164.

68. Martin AR, Kanai M, Kamatani Y, Okada Y, Neale BM, Daly MJ: Clinical use of current polygenic risk scores may exacerbate health disparities. Nat Genet 2019, 51(4):584–591.

69. Liang Y, Pividori M, Manichaikul A, Palmer AA, Cox NJ, Wheeler HE, Im HK: Polygenic transcriptome risk scores (PTRS) can improve portability of polygenic risk scores across ancestries. Genome Biol 2022, 23(1):23.

70. Zeng J, Xue A, Jiang L, Lloyd-Jones LR, Wu Y, Wang H, Zheng Z, Yengo L, Kemper KE, Goddard ME et al: Widespread signatures of natural selection across human complex traits and functional genomic categories. Nat Commun 2021, 12(1):1164.

